# Variant Classification of a Non-Protein Coding Gene: ACMG/AMP Specifications for *RMRP* from the Clinical Genome Resource Severe Combined Immunodeficiency Disease Variant Curation Expert Panel

**DOI:** 10.64898/2026.11.02.26366506

**Authors:** Vanessa C. Jacovas, Yasemin Demirkol, Alejandro Nieto-Patlan, Shiloh Martin, Shannon McNulty, Eric J. Allenspach, Svetlana Vakkilainen, Luigi D. Notarangelo, Jennifer M. Puck, Jolan Walter, Alice Y. Chan, Morton J. Cowan, Michael S. Hershfield, Nicolai S. C. van Oers, James Verbsky, Brian H.Y. Chung, Rajarshi Ghosh, Xi Luo, Megan Piazza, Katharina V. Schulze, Theru A. Sivakumaran, Bo Yuan, Britt A. Johnson, Ivan K. Chinn, Severe Combined Immunodeficiency Disease Variant Curation Expert Panel

## Abstract

**Purpose:** This study, conducted by the Clinical Genome Resource Severe Combined Immunodeficiency Disease Variant Curation Expert Panel (ClinGen SCID-VCEP), aimed to adapt the American College of Medical Genetics and Genomics/Association for Molecular Pathology (ACMG/AMP) framework for germline variant interpretation in *RMRP*. Unlike the protein-coding genes for which the framework was primarily developed, *RMRP* encodes a non-protein-coding RNA. Biallelic pathogenic variants in *RMRP* cause cartilage–hair hypoplasia, which can include severe combined immunodeficiency disease and other immune abnormalities.

**Methods:** An *RMRP*-focused SCID-VCEP subgroup reviewed all 28 ACMG/AMP evidence criteria using population databases, published clinical and experimental data, computational predictions, and expert consensus. Each criterion was retained, modified, or designated as not applicable based on *RMRP* biology and disease mechanisms. The resulting specifications were pilot tested using 27 variants selected from ClinVar and research laboratory data.

**Results:** Fifteen ACMG/AMP criteria were determined to be applicable to *RMRP*, while 13 were considered not applicable. Several criteria were modified to evaluate nucleotide-level effects on transcriptional regulation, critical promoter regions, and RNA secondary structure. The specifications also incorporated *RMRP*-specific recommendations for population frequency, functional evidence, variant phase, segregation, and phenotype. Application of the specifications classified 6 variants as pathogenic, 10 as likely pathogenic, 7 as variants of uncertain significance, and 4 as benign. Previously pathogenic or likely pathogenic variants remained within the pathogenic spectrum, and all previously benign variants remained benign. Variants with conflicting ClinVar interpretations were resolved as either likely pathogenic or uncertain. PM1, PM3, and PP4 were the primary criteria supporting pathogenic classifications.

**Conclusion:** The SCID-VCEP specifications provide a standardized approach to the classification of germline *RMRP* variants in cartilage–hair hypoplasia and demonstrate how the ACMG/AMP framework can be adapted for a disease-associated non-protein-coding RNA gene. These specifications support the development of variant interpretation frameworks for other clinically relevant non-protein-coding genes.

## Introduction

Severe Combined Immunodeficiency Disease (SCID) is a life-threatening inborn error of immunity characterized by profound defects in both cellular and humoral immune function due to impaired T- and B-lymphocyte development and function. Clinically, SCID presents in early infancy with severe, recurrent, and opportunistic infections, persistent viral or fungal disease, and failure to thrive.^1^ Without early diagnosis and treatment, the condition is typically fatal within the first year of life.^2,3^ With an estimated incidence of approximately 1 in 50,000 live births in North America, timely recognition of SCID is essential.^4^ The implementation of population-based newborn screening programs has enabled early detection, allowing prompt initiation of curative therapies, such as hematopoietic stem cell transplantation or gene therapy, and significantly improving survival outcomes.^2,3^

Due to the critical role of the RNase mitochondrial RNA processing (MRP) complex in rapidly dividing immune cells, pathogenic variants in its RNA component-encoding gene, *RMRP*, can result in SCID in newborns.^1^ *RMRP* (historically NR_003051.3), first identified in the early 1990s through studies of ribosomal RNA processing, encodes the RNA component of the MRP complex and represents one of the first disease-causing non-protein coding RNA genes described in humans.^5,6^ The ribonucleoprotein endoribonuclease participates in multiple nuclear and mitochondrial processes, including ribosomal RNA maturation, cell cycle regulation, and mitochondrial DNA replication.^6-9^ It is therefore essential for normal cellular proliferation, particularly in rapidly dividing tissues. Biallelic germline pathogenic loss-of-function variants in *RMRP* cause cartilage–hair hypoplasia (CHH; OMIM #250250, MONDO:0009595), a rare condition characterized by a wide range of clinical features, including metaphyseal skeletal abnormalities, disproportionate short stature, joint hypermobility, fine and lightly pigmented sparse hair, gastrointestinal issues, anemia, combined immunodeficiency, and increased risk for malignancies such as lymphoma.^6,10-12^

The Clinical Genome Resource (ClinGen) SCID Variant Curation Expert Panel (VCEP) previously established gene-specific American College of Medical Genetics and Genomics (ACMG)/Association for Molecular Pathology (AMP) variant interpretation rules for seven protein-coding genes most frequently implicated in SCID.^13^ This current study represents a focused extension of that framework to *RMRP*. Adapting the ACMG/AMP variant interpretation framework to a non-protein coding RNA gene presented unique conceptual and technical challenges, as many standard evidence categories are focused upon protein-coding changes and rely on predicted effects on amino acid sequence, protein structure, or protein function. For *RMRP*, pathogenicity arises from disruptions to RNA structure, expression levels, or ribonucleoprotein complex function rather than a single altered protein product. This gene product difference mandates careful reassessment of how existing ACMG/AMP criteria can be applied, modified, or constrained for this gene. In this study, SCID-VCEP–approved ACMG/AMP specifications tailored to *RMRP* for CHH were established, representing the first systematic attempt to adapt the widely used variant classification framework to a disease-causing non-protein coding RNA gene and providing a foundation for future variant interpretation efforts in non-coding regions of the genome.

## Materials and Methods

### Formation of an RMRP Expert Subgroup within the SCID-VCEP

The SCID-VCEP established a dedicated *RMRP* working subgroup that met monthly. This subgroup included experts with relevant expertise in *RMRP* biology and CHH, immunodeficiency, and clinical genetics. Variant classifications were performed with discussion within the *RMRP* subgroup and subsequently reviewed by the full SCID-VCEP.

### Modification of ACMG/AMP Criteria for the Non-Protein Coding RMRP Gene

Adaptation of the ACMG/AMP sequence variant interpretation guidelines^14^ towards *RMRP* was systematically performed through regular subgroup meetings. Each ACMG/AMP criterion was carefully reviewed and modified to accommodate the unique biological characteristics of *RMRP* as a non-protein coding gene, for which conventional protein-based evidence categories are not applicable. As a result, gene-centric specifications were established that emphasized RNA structure, functional studies, segregation data, and clinical phenotype rather than protein impact.

### Validation and Pilot Testing

The adapted specifications for CHH were subsequently evaluated through pilot testing, in which 27 *RMRP* variants were curated by trained biocurators using available published and internal laboratory evidence. Of note, many published studies refer to legacy transcripts NR_003051.1 and NR_003051.2, which differ by a single ‘t’ to ‘c’ change at n.53 and which were superseded by NR_003051.3 in 2010. In this report, all variant positions are given relative to NR_003051.3, since the VCEP developed and tested the rules specifications using this widely used transcript. NR_003051.3 was recently changed to NR_003051.4 by Matched Annotation from the National Center for Biotechnology Information and European Molecular Biology Laboratory’s European Bioinformatics Institute (MANE). The MANE transcript is shuffled 1 nucleotide to the left compared to NR_003051.3 (starting with ‘tggttcgtgc…’ for NR_003051.4 vs. ‘ggttcgtgc…’ for NR_003051.3) and 2 nucleotides to the left compared to NR_003051.1 and NR003051.2 (which start with ‘gttcgtgc…’). Due to multiple transcripts, positions may appear to differ by 1 or 2 nucleotides online in some places. Variants are linked to an identical ClinVar ID across all transcripts to ensure consistency. Consensus between the SCID-VCEP members was obtained to secure the final classification of the variants (benign, likely benign, variant of uncertain significance, likely pathogenic or pathogenic) based on the ACMG/AMP evidence code combinations. The refined *RMRP*-specific classification criteria were reviewed and endorsed by the ClinGen Sequence Variant Interpretation Working Group and are publicly available via the ClinGen Criteria Specification Registry, where they are updated when new evidence emerges.

## Results

Of the 28 original ACMG/AMP criteria, 15 were defined as applicable for *RMRP*: BA1, BS1, BS2, BS4, PP1, PP3, PP4, PM1, PM2, PM3, PM4, PM6, PS1, PS2, and PS3. Given the non-protein coding nature of *RMRP*, multiple ACMG/AMP criteria were defined as not applicable, as they rely on protein sequence, protein structure, or amino acid conservation (BP1, BP7, PP2, PM5, PVS1); assume recurrent mutational patterns or informative repetitive sequence contexts (BP3); are incompatible with the established autosomal recessive inheritance of *RMRP*-associated disease (PS4); or depend on external variant classifications discouraged by ClinGen recommendations (BP6, PP5). Overall, 13 were defined as not applicable for *RMRP*: BP1, BP2, BP3, BP4, BP5, BP6, BP7, BS3, PP2, PP5, PM5, PS4, and PVS1. This section describes the applicability, modification, and implementation of ACMG/AMP criteria as specified for *RMRP*.

### Modification of the ACMG/AMP criteria based on specific classes of criteria

#### Variant frequency and use of control populations (BA1, BS1, BS2, PM2, and PS4)

The allele frequency calculator (http://cardiodb.org/allelefrequencyapp) was used to derive the minor allele frequency (MAF) thresholds for BA1, BS1 and PM2. Thresholds were calculated using disease prevalence, allelic heterogeneity, maximum genetic contribution (genetic heterogeneity), and penetrance. For *RMRP*, genetic heterogeneity was set to 0.04 (4%), based on the contribution of *RMRP* variants to total SCID cases reported in North America.^1,15^ For BA1 (allele frequency greater than expected for disorder, Stand-alone), penetrance was set to 50%, and disease prevalence was set at 1 in 5,000, representing the highest reported prevalence of SCID^16^ and a conservative approach that yielded a stringent BA1 threshold of 0.00400. For BS1 (allele frequency greater than expected for disorder), penetrance was set to 100%, and the disease prevalence was set at 1 in 50,000, corresponding to the general number for the North American population^17,18^ and resulting in a BS1 threshold of 0.00089.

PM2 (absent from population databases) provides evidence for variant pathogenicity based on absent or extremely rare minor allele frequency. For this criterion, calculations were performed by reducing the allelic heterogeneity parameter, as previously implemented in ClinGen SCID-VCEP gene specifications,^13^ and the threshold was derived at 0.0000447. PM2 was also applied at the Supporting level based on ClinGen recommendations (https://clinicalgenome.org/docs/pm2-recommendation-for-absence-rarity/). Furthermore, to apply PM2, the variant cannot be observed in the homozygous state within any population.

The BS2 code (observed in a healthy adult individual for a recessive disorder with full penetrance at an early age) assesses whether a variant is found in unaffected adult individuals with early-onset, highly penetrant conditions. Observing a homozygous adult without clinical features of the disease supports a benign classification. For *RMRP*, experts determined that BS2 can be applied at the Supporting level when at least two unaffected homozygous adults are reported in the literature or 2 homozygotes are identified in population databases, such as gnomAD. For a strong-level application, the variant must be observed in at least 3 homozygotes in population databases or homozygous unaffected adults.

PS4 (prevalence of the variant in affected individuals is significantly increased compared with the prevalence in controls) was not applied because evidence from enumeration of affected individuals is assessed using PM3 rather than case-control enrichment analysis for autosomal recessive disorders.

#### Functional evidence (BS3, PS3, and PP2)

For PS3 (well established *in vitro* or *in vivo* functional studies show damaging effect on protein function or splicing), the SCID-VCEP selected specific assays that have demonstrated benign or damaging effects across multiple *RMRP* variants. This selection was based on a comprehensive review of literature and expert opinions from SCID-VCEP members. It was determined that consideration of any animal model representing the variant of interest would be reviewed by the SCID-VCEP on a case-by-case manner. *In vitro* assays accepted by the VCEP included an endonucleolytic cleavage activity assay^9,19^ or luciferase reporter assay^20^. In terms of endonucleolytic cleavage, the VCEP recognized that reduced processing of both rRNA and mRNA is specific for CHH, whereas impaired rRNA processing without defective mRNA cleavage correlates instead with anauxetic dysplasia (OMIM #607095, MONDO: 0011773).^9^ As a result, mRNA cleavage activity less than 0.9 is required to apply PS3. Due to lack of sufficient benign and pathogenic controls in isolated assays, PS3 strength for *in vitro* evidence was downgraded to Supporting.^21^ Table S1 compiles all the functional assays considered for *RMRP* variant classification.

For BS3, the VCEP felt that insufficient methodology exists to confirm benignity of potential variants, as all functional evidence depends on specific designs, models, or interpretations. As a result, this code was considered not applicable to avoid uncertainty.

Finally, the PP2 criterion (missense variant in a gene that has a low rate of benign missense variation and where missense variants are a common mechanism of disease) was also deemed inapplicable because *RMRP* is a non-protein coding gene and therefore does not contain missense variants. Consequently, gene-level missense constraint metrics, such as the gnomAD missense Z-score used to support PP2, do not apply to *RMRP*.

#### Critical domain (PM1)

PM1 (mutational hot spots and well-established functional domains) is defined as applicable at the Strong level for *RMRP.* This evidence includes insertions or duplications occurring within the core promoter region, specifically between the TATA box (n.-32 to n.-24) and downstream to the transcription start site, up to position n.4. Experimental studies have shown that sequence alterations in this promoter region significantly impair transcriptional activity, supporting its role as a key functional regulatory element.^5,22-24^ To apply PM1, the variant must be sufficiently rare in the general population. While such variants are not required to meet PM2 criteria, they must not exceed BS1 or BA1 frequency thresholds unless a founder effect is suspected. If a variant exceeds BS1 or BA1 thresholds due to a suspected founder effect, its applicability for PM1 will be evaluated on a case-by-case basis by the SCID-VCEP.

#### Computational and predictive data (BP1, BP3, BP4, BP7, PP3, PM4, PM5, PS1, and PVS1)

Because *RMRP* is a non-protein coding gene, the SCID-VCEP reformulated several ACMG/AMP criteria to ensure their applicability. When possible, codes which traditionally rely on amino acid–level changes were adapted to evaluate pathogenicity at the nucleotide level.

The PM4 code is traditionally applied as pathogenic evidence for variants that alter protein length through insertions or deletions. This criterion was modified to capture pathogenic mechanisms relevant to transcriptional regulation. Specifically, PM4 was adapted to specify pathogenic evidence for insertions that increase the distance between the TATA box (positions −32 to −24) and the transcription start site (positions +1 to +4) *by six or more* nucleotides, a configuration expected to disrupt normal transcriptional initiation.^6^ PM4 is distinguished from PM1 by the notion that PM1 only addresses the location of a variant relative to the TATA box, not its impact on the distance between the TATA box and the start site.

PS1 is traditionally applied as pathogenic evidence when a variant results in the same amino acid change as a previously established pathogenic variant. For *RMRP*, the SCID-VCEP modified this criterion by downgrading its strength and adapting its application to nucleotide-level changes. Under this specification, PS1 is applied at a Supporting level when a different nucleotide substitution occurs at the same genomic position as a variant previously classified as pathogenic by the SCID-VCEP. This evidence is not applied if a benign variant has also been classified at the same nucleotide position.

Some codes could not be adapted in similar fashion because they rely heavily on protein translation: BP1 (missense in a gene where only truncation causes disease), BP3 (in-frame indels in repetitive region without known function), BP7 (silent nucleotide change), PM5 (missense change at the same amino acid residue as a different pathogenic missense change), and PVS1 (predicted null variant). They were therefore considered not applicable and were excluded from variant classification.

The SCID-VCEP also agreed to allow Supporting pathogenic evidence from *in silico* analyses when variants are predicted to disrupt the RNA secondary structure. The RNAsnp tool developed by the Center for Non-Coding RNA in Technology and Health was approved due to its ability to consider secondary RNA structure.^25,26^ Only variants located within the transcribed region of *RMRP* were eligible for PP3 consideration; variants located in the promoter region were not accepted (Mode: 1, Folding window: 100 nt, Measure: “Distance”, Minimum length: 50, Cut-off for base pair probabilities: 0.01). Consistent with prior studies, a p-value of less than 0.1 was used to define statistical significance for predicted structural effects.^25^ On the other hand, in the absence of calibration analyses, the VCEP elected against use of *in silico* predictions for benign evidence (BP4).

#### Case/segregation codes (BS4, PP1, PM6, PS2, and PM3)

For PP1 (cosegregation with disease in affected family members), cosegregation evidence is evaluated using the number of informative affected and unaffected family members together with the calculated logarithm of the odds (LOD) score to determine the strength of evidence. PP1 is applied at a supporting level for LOD scores ≥0.6 and <1.2, at a moderate level for scores ≥1.2 and <1.5, and at a strong level for scores ≥1.5.^27^ The BS4 criterion (lack of segregation in affected family members) is applied without additional gene-specific modifications and used when an individual with an *RMRP*-associated phenotype does not carry the variant under evaluation.

Although rare, *de novo RMRP* variants have been reported in the literature associated with CHH.^22^ To address these cases, PM6 *de novo* (maternity and paternity not confirmed) and PS2 *de novo* (maternity and paternity confirmed) criteria were defined as applicable following standard ClinGen recommendations (Table S2).^28^ For application of PM6 and PS2, the strength of evidence is determined based on the cumulative point total across all probands, where a total of 0.5 points corresponds to Supporting evidence, 1 point to Moderate, 2 points to Strong, and 4 points to Very Strong. To define the phenotypic consistency (“phenotype highly specific for gene”), the proband must meet PP4_Moderate criteria. For “phenotype consistent with the gene but not highly specific,” the proband must meet PP4. For “phenotype consistent with gene but not highly specific and high genetic heterogeneity,” the proband must have been asserted to have a CHH phenotype but lack sufficient description to meet PP4 criteria.

PM3 evidence (variant detected *in trans* with a pathogenic variant) followed ClinGen guidance for proband-based point assignment (https://clinicalgenome.org/working-groups/sequence-variant-interpretation). Higher point values are assigned when a pathogenic or likely pathogenic *RMRP* variant is conclusively demonstrated to be *in trans* with another pathogenic variant through parental testing or cloning-based approaches. Lower point values are applied to configurations in which the *in trans* status is inferred but not experimentally confirmed, as well as to homozygous occurrences. Use of PM3 requires that both variants in compound heterozygous cases, or the single variant in homozygous individuals, meet the allele frequency threshold defined by PM2. For *RMRP* variants suspected to represent founder alleles and therefore observed at frequencies exceeding the PM2 threshold, the appropriateness of applying PM3 is evaluated on a case-by-case basis by the SCID-VCEP.

#### Phenotype (PP4)

The SCID-VCEP developed a phenotype scoring system to determine PP4 strength (Table 1). SCID was defined using Primary Immune Deficiency Treatment Consortium (PIDTC) scoring criteria.^1^ At the same time, the VCEP recognized that SCID comprises only one of various potential manifestations of CHH. Other key features include skeletal dysplasia, hypotrichosis, macrocytic or hypoplastic anemia, and Hirschsprung disease or congenital megacolon.^6,10-12^ Importantly, the cumulative presence of these characteristics was granted increased strength for PP4 by the VCEP from Supporting up to Moderate.

**Table 1.** PP4 Phenotype Scoring Criteria for *RMRP*.

| Clinical features | Points awarded |
|---|---|
| Diagnostic criteria met for SCID/Leaky SCID/Omenn syndrome<sup>a</sup> | 1 point |
| SCID gene panel or exome/genome sequencing conducted<sup>b</sup> | 0.5 points |
| Family history of SCID | 0.5 points |
| Family history of CHH | 1 point |
| Metaphyseal dysplasia (disproportionate short stature + radiographic evidence) | 1 point |
| Skeletal dysplasia gene panel or exome/genome sequencing conducted with no alternative genetic diagnosis | 1 point |
| Hypotrichosis | 0.5 points |
| Macrocytic, hypoplastic anemia | 0.25 points |
| Hirschsprung disease or congenital megacolon | 0.25 points |
| T-cell lymphopenia<sup>c</sup> | 0.5 points |

<sup>a</sup> The diagnostic criteria should follow the PIDTC 2022 specification (PMID: 36456361)
<sup>b</sup> Only applicable if genetic testing did not provide an alternative genetic explanation for SCID/Leaky SCID/Omenn syndrome phenotype
<sup>c</sup> If maternal T cells are present, the T lymphocyte profile is still considered to be T- (autologous T cells are absent). Allocate 0.25 points for T-cell lymphopenia only in cases where the SCID diagnostic criteria are not applied. It is important to note that if the SCID diagnostic criteria were applied, the points for the T-B+NK+ lymphocyte subset profile cannot be considered.

#### Combining Rules

In addition to specifying individual ACMG/AMP criteria, the SCID-VCEP modified two ACMG/AMP combining rules for *RMRP* variant classification. The combination of 1 (one) Very Strong pathogenic criterion and one (1) Supporting pathogenic criterion was defined as sufficient for Likely Pathogenic classification. Likewise, 1 (one) strong benign criterion was considered sufficient for Likely Benign classification. All remaining rules followed the ACMG/AMP framework.

### Performance of the adapted RMRP ACMG/AMP specifications in variant classification

The adapted *RMRP* (NR_003051.3) specifications for CHH were tested using a pilot set of 27 variants (Table 2). The variants were selected from ClinVar and research laboratory data. They included variants with previous assertions of benign/likely benign (B/LB), pathogenic/likely pathogenic (P/LP), variant of uncertain significance (VUS), and variants with absent or conflicting interpretations. The selection comprised very well-established *RMRP* pathogenic variants as well as variants located in both the transcript and promoter regions to allow for a comprehensive assessment of specification performance. Case segregation and functional evidence were gathered from internal data and available published literature.

**Table 2.**
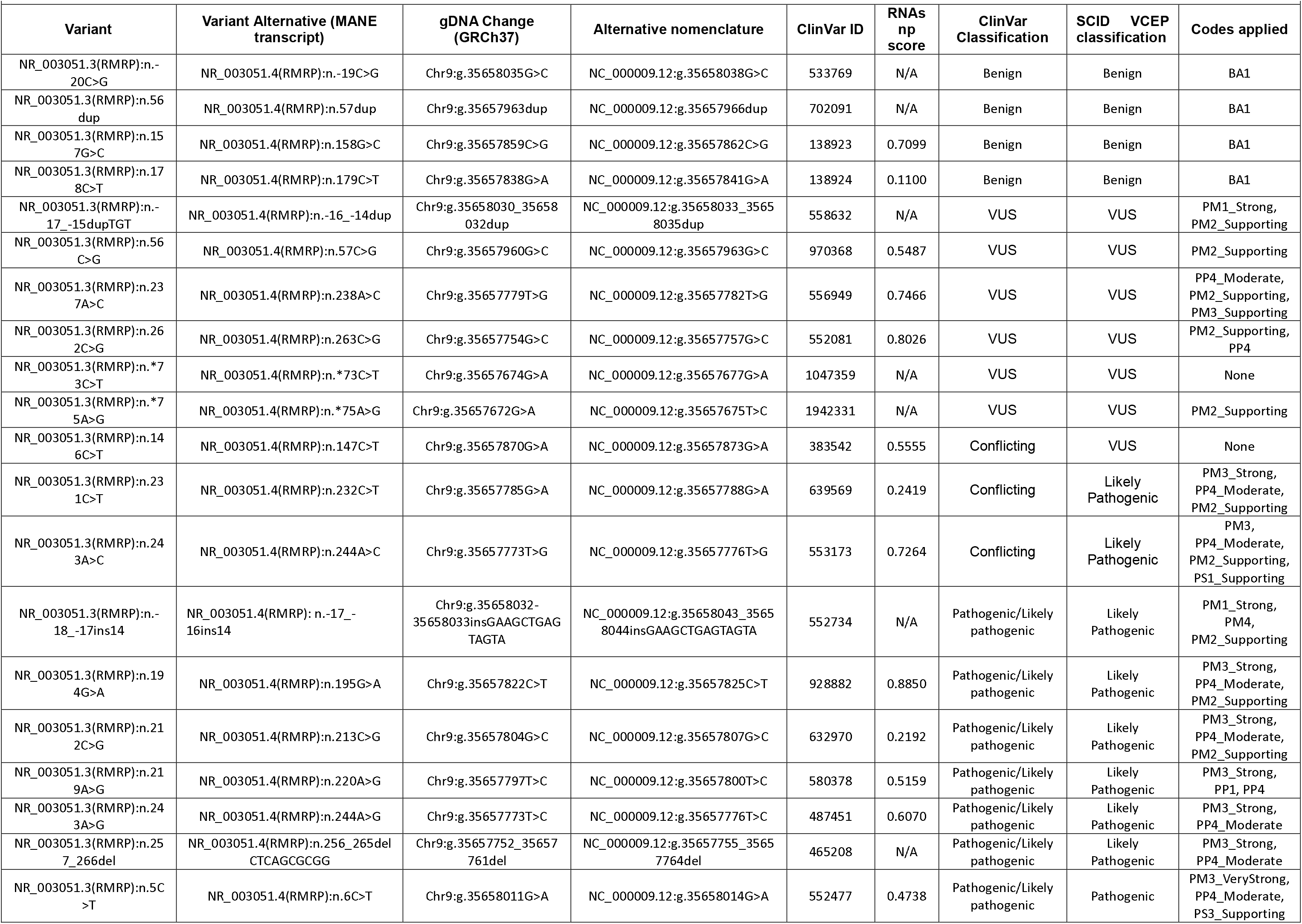
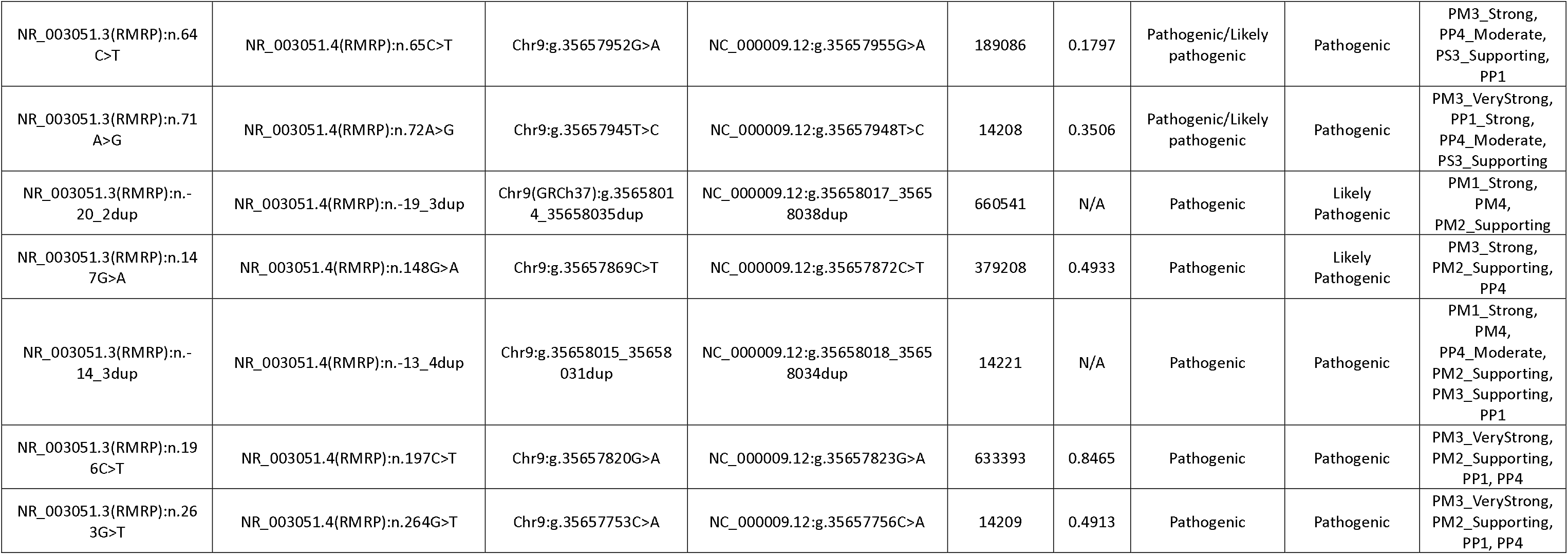
*RMRP* variants classified for cartilage-hair hypoplasia during pilot testing.

| Variant | Variant Alternative (MANE transcript) | gDNA Change (GRCh37) | Alternative nomenclature | ClinVar ID | RNAsnp score | ClinVar Classification | SCID VCEP classification | Codes applied |
|---|---|---|---|---|---|---|---|---|
| NR_003051.3(RMRP):n.-20C>G | NR_003051.4(RMRP):n.-19C>G | Chr9:g.35658035G>C | NC_000009.12:g.35658038G>C | 533769 | N/A | Benign | Benign | BA1 |
| NR_003051.3(RMRP):n.56dup | NR_003051.4(RMRP):n.57dup | Chr9:g.35657963dup | NC_000009.12:g.35657966dup | 702091 | N/A | Benign | Benign | BA1 |
| NR_003051.3(RMRP):n.157G>C | NR_003051.4(RMRP):n.158G>C | Chr9:g.35657859C>G | NC_000009.12:g.35657862C>G | 138923 | 0.7099 | Benign | Benign | BA1 |
| NR_003051.3(RMRP):n.178C>T | NR_003051.4(RMRP):n.179C>T | Chr9:g.35657838G>A | NC_000009.12:g.35657841G>A | 138924 | 0.1100 | Benign | Benign | BA1 |
| NR_003051.3(RMRP):n.-17_-15dupTGT | NR_003051.4(RMRP):n.-16_-14dup | Chr9:g.35658030_35658032dup | NC_000009.12:g.35658033_35658035dup | 558632 | N/A | VUS | VUS | PM1_Strong, PM2_Supporting |
| NR_003051.3(RMRP):n.56C>G | NR_003051.4(RMRP):n.57C>G | Chr9:g.35657960G>C | NC_000009.12:g.35657963G>C | 970368 | 0.5487 | VUS | VUS | PM2_Supporting |
| NR_003051.3(RMRP):n.237A>C | NR_003051.4(RMRP):n.238A>C | Chr9:g.35657779T>G | NC_000009.12:g.35657782T>G | 556949 | 0.7466 | VUS | VUS | PP4_Moderate, PM2_Supporting, PM3_Supporting |
| NR_003051.3(RMRP):n.262C>G | NR_003051.4(RMRP):n.263C>G | Chr9:g.35657754G>C | NC_000009.12:g.35657757G>C | 552081 | 0.8026 | VUS | VUS | PM2_Supporting, PP4 |
| NR_003051.3(RMRP):n.*73C>T | NR_003051.4(RMRP):n.*73C>T | Chr9:g.35657674G>A | NC_000009.12:g.35657677G>A | 1047359 | N/A | VUS | VUS | None |
| NR_003051.3(RMRP):n.*75A>G | NR_003051.4(RMRP):n.*75A>G | Chr9:g.35657672G>A | NC_000009.12:g.35657675T>C | 1942331 | N/A | VUS | VUS | PM2_Supporting |
| NR_003051.3(RMRP):n.146C>T | NR_003051.4(RMRP):n.147C>T | Chr9:g.35657870G>A | NC_000009.12:g.35657873G>A | 383542 | 0.5555 | Conflicting | VUS | None |
| NR_003051.3(RMRP):n.231C>T | NR_003051.4(RMRP):n.232C>T | Chr9:g.35657785G>A | NC_000009.12:g.35657788G>A | 639569 | 0.2419 | Conflicting | Likely Pathogenic | PM3_Strong, PP4_Moderate, PM2_Supporting |
| NR_003051.3(RMRP):n.243A>C | NR_003051.4(RMRP):n.244A>C | Chr9:g.35657773T>G | NC_000009.12:g.35657776T>G | 553173 | 0.7264 | Conflicting | Likely Pathogenic | PM3, PP4_Moderate, PM2_Supporting, PS1_Supporting |
| NR_003051.3(RMRP):n.-18_-17ins14 | NR_003051.4(RMRP): n.-17_-16ins14 | Chr9:g.35658032-35658033insGAAGCTGAGTAGTA | NC_000009.12:g.35658043_35658044insGAAGCTGAGTAGTA | 552734 | N/A | Pathogenic/Likely pathogenic | Likely Pathogenic | PM1_Strong, PM4, PM2_Supporting |
| NR_003051.3(RMRP):n.194G>A | NR_003051.4(RMRP):n.195G>A | Chr9:g.35657822C>T | NC_000009.12:g.35657825C>T | 928882 | 0.8850 | Pathogenic/Likely pathogenic | Likely Pathogenic | PM3_Strong, PP4_Moderate, PM2_Supporting |
| NR_003051.3(RMRP):n.212C>G | NR_003051.4(RMRP):n.213C>G | Chr9:g.35657804G>C | NC_000009.12:g.35657807G>C | 632970 | 0.2192 | Pathogenic/Likely pathogenic | Likely Pathogenic | PM3_Strong, PP4_Moderate, PM2_Supporting |
| NR_003051.3(RMRP):n.219A>G | NR_003051.4(RMRP):n.220A>G | Chr9:g.35657797T>C | NC_000009.12:g.35657800T>C | 580378 | 0.5159 | Pathogenic/Likely pathogenic | Likely Pathogenic | PM3_Strong, PP1, PP4 |
| NR_003051.3(RMRP):n.243A>G | NR_003051.4(RMRP):n.244A>G | Chr9:g.35657773T>C | NC_000009.12:g.35657776T>C | 487451 | 0.6070 | Pathogenic/Likely pathogenic | Likely Pathogenic | PM3_Strong, PP4_Moderate |
| NR_003051.3(RMRP):n.257_266del | NR_003051.4(RMRP):n.256_265delCTCAGCGCGG | Chr9:g.35657752_35657761del | NC_000009.12:g.35657755_35657764del | 465208 | N/A | Pathogenic/Likely pathogenic | Likely Pathogenic | PM3_Strong, PP4_Moderate |
| NR_003051.3(RMRP):n.5C>T | NR_003051.4(RMRP):n.6C>T | Chr9:g.35658011G>A | NC_000009.12:g.35658014G>A | 552477 | 0.4738 | Pathogenic/Likely pathogenic | Pathogenic | PM3_VeryStrong, PP4_Moderate, PS3_Supporting |
| NR_003051.3(RMRP):n.64C>T | NR_003051.4(RMRP):n.65C>T | Chr9:g.35657952G>A | NC_000009.12:g.35657955G>A | 189086 | 0.1797 | Pathogenic/Likely pathogenic | Pathogenic | PM3_Strong, PP4_Moderate, PS3_Supporting, PP1 |
| NR_003051.3(RMRP):n.71A>G | NR_003051.4(RMRP):n.72A>G | Chr9:g.35657945T>C | NC_000009.12:g.35657948T>C | 14208 | 0.3506 | Pathogenic/Likely pathogenic | Pathogenic | PM3_VeryStrong, PP1_Strong, PP4_Moderate, PS3_Supporting |
| NR_003051.3(RMRP):n.-20_2dup | NR_003051.4(RMRP):n.-19_3dup | Chr9(GRCh37):g.35658014_35658035dup | NC_000009.12:g.35658017_35658038dup | 660541 | N/A | Pathogenic | Likely Pathogenic | PM1_Strong, PM4, PM2_Supporting |
| NR_003051.3(RMRP):n.147G>A | NR_003051.4(RMRP):n.148G>A | Chr9:g.35657869C>T | NC_000009.12:g.35657872C>T | 379208 | 0.4933 | Pathogenic | Likely Pathogenic | PM3_Strong, PM2_Supporting, PP4 |
| NR_003051.3(RMRP):n.-14_3dup | NR_003051.4(RMRP):n.-13_4dup | Chr9:g.35658015_35658031dup | NC_000009.12:g.35658018_35658034dup | 14221 | N/A | Pathogenic | Pathogenic | PM1_Strong, PM4, PP4_Moderate, PM2_Supporting, PM3_Supporting, PP1 |
| NR_003051.3(RMRP):n.196C>T | NR_003051.4(RMRP):n.197C>T | Chr9:g.35657820G>A | NC_000009.12:g.35657823G>A | 633393 | 0.8465 | Pathogenic | Pathogenic | PM3_VeryStrong, PM2_Supporting, PP1, PP4 |
| NR_003051.3(RMRP):n.263G>T | NR_003051.4(RMRP):n.264G>T | Chr9:g.35657753C>A | NC_000009.12:g.35657756C>A | 14209 | 0.4913 | Pathogenic | Pathogenic | PM3_VeryStrong, PM2_Supporting, PP1, PP4 |

The *RMRP* specifications facilitated classification of the 27 *RMRP* variants: 6 as P, 10 as LP, 7 as VUS, and 4 as benign. Changes from previous ClinVar classifications are represented in Figure 1. No B, LP, or P variants were reclassified as VUS. All P/LP classifications emerged as either P or LP. Notably, all conflicting classifications were resolved as either VUS or LP. Every VCEP-classified P and LP variant except two (n.-20_2dup and n.-14_3dup) required use of PM3 at default strength or higher. The 2 exceptions instead used PM1_Strong to achieve their classifications. PP4 was necessary to upgrade variants from VUS to LP in 8 of 10 (80%) variants and from LP to P in 3 of 6 (50%) variants. On the other hand, all variants classified as B used BA1. None of the 7 VCEP-determined VUS met BS1 or BS2. Notably, only 2 had sufficient phenotypic information available to apply PP4, none had any functional data support, and none met criteria to apply PP3. The classifications were approved by the general SCID-VCEP and deposited in the ClinGen Evidence Repository and ClinVar.

**Figure 1.**
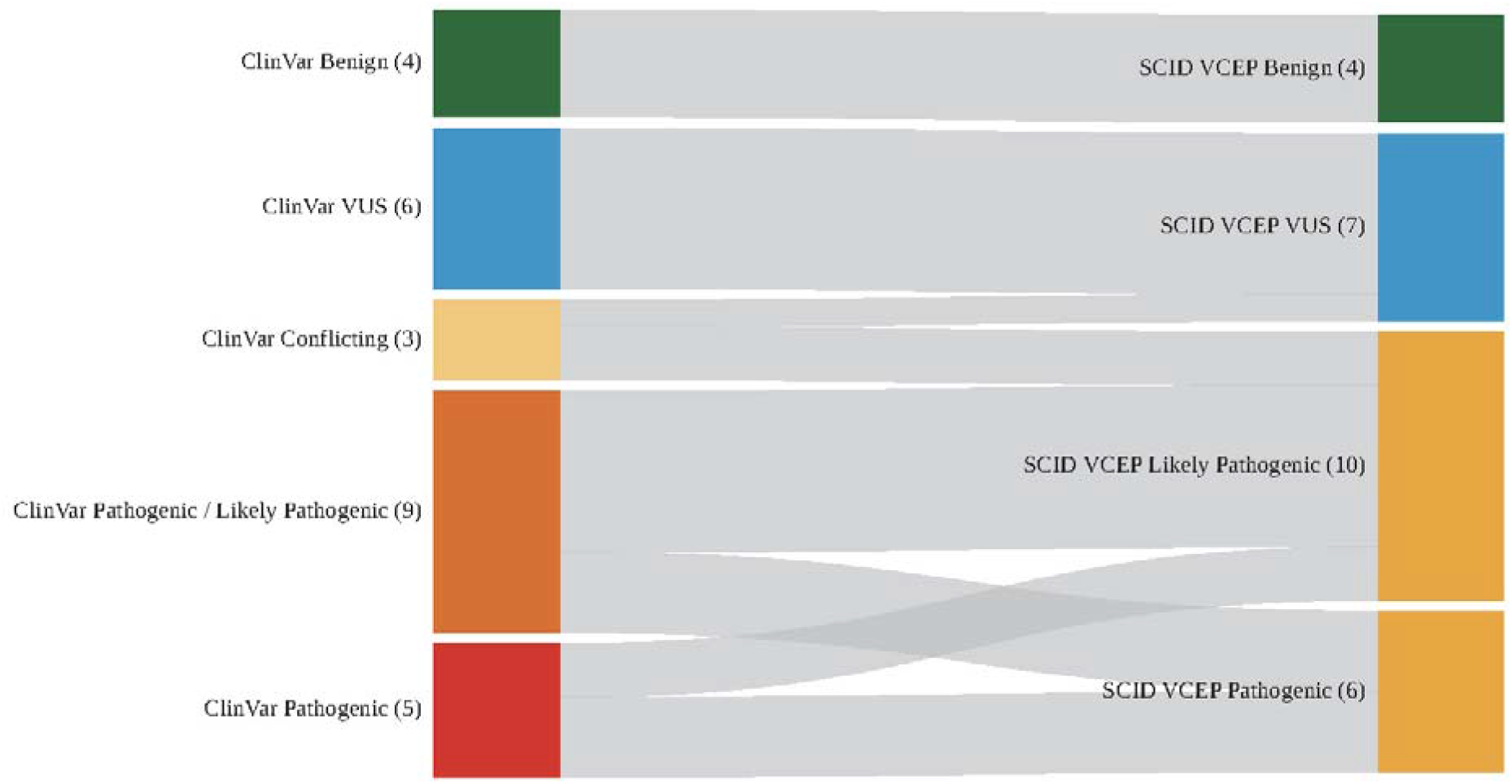
Classifications of the 27 selected *RMRP* variants by the original ClinVar assertion (left) and the SCID VCEP (right). The raw numbers in parentheses represent the total number of variants in each classification category. Conflicting classifications indicate variants for which ClinVar contained assertions from multiple submitters in conflicting or potentially conflicting pathogenicity classification categories (*e.g.*, likely benign and VUS or VUS and likely pathogenic). Differing assertions within the same pathogenicity spectrum (*i.e.*, likely pathogenic and pathogenic or benign and likely benign) are not considered conflicting by ClinVar and are grouped separately. SCID VCEP, Severe Combined Immunodeficiency Disease Variant Curation Expert Panel; VUS, variant of uncertain significance.

## Discussion

The ClinGen SCID-VCEP has successfully piloted classification specifications for evaluating the pathogenicity of *RMRP* variants in CHH, representing a landmark achievement for application of the ACMG/AMP rules framework toward a non-protein coding gene. This process required expert knowledge concerning RNase MRP complex biology, transcription of the RNA component, and human disease resulting from its loss. Using this information, ACMG/AMP evidence codes were adapted to fit *RMRP* as best possible. Ultimately, from 28 evidence codes, 15 were deemed applicable and 13 were considered not applicable for *RMRP* variant classification. The utility of these *RMRP*-specific rules specifications for CHH was verified during pilot testing. ClinVar-asserted B variants remained B, and pathogenic spectrum variants remained either P or LP. The specifications also successfully enabled appropriate classification of variants with conflicting classifications.

Key evidence codes that drove pathogenic classification included the following: PM1, PM3, and PP4. Upgraded strengths for PM1 and PP4 reflect expert knowledge of *RMRP* among the VCEP subgroup members. For instance, the decision to allow PM1 strength to be increased to Strong incorporates knowledge that increased distance between the TATA box and transcription initiation site has been well-established as a critical cause of decreased *RMRP* RNA levels. Expert understanding of the clinical features associated with *RMRP* deficiency also allowed PP4 to be appropriately advanced up to Moderate strength. Meanwhile, the utility of PM3 reflects the notion that CHH and SCID are both highly rare conditions, so each identified case provides valuable evidence for pathogenicity.

The rules development and pilot testing process also revealed specific limitations that will need to continue to be addressed, especially since all ClinVar-asserted VUS remained as VUS. First, the heavy reliance on 3 codes for pathogenic classification often meant that variants with insufficient data to apply these codes at higher levels of strength remanded VCEP classifications to VUS. In particular, the inability to apply or upgrade PM3 and PP4 highlighted either a lack of published cases (PM3) or poverty of phenotypic information whenever cases were reported (PP4). When the VCEP was able to find additional unpublished cases by communicating with ClinVar submitters or other sources, these entities often represented laboratories that did not have requisite PP4 data. Second, the VCEP activities also identified a lack of functional testing data to support use of PS3 and BS3. In fact, the VCEP was able to apply PS3_Supporting to only 3 variants, and for 2 of the variants, Pathogenic classification was achievable without the code. Finally, the lack of utility for PP3 illustrated absence of computational prediction algorithm calibration for *RMRP*. The challenge faced by the VCEP involved the concept that almost all such algorithms have been trained upon effect on protein expression, structure, or function. At the time of specifications development, RNAsnp represented one of few algorithms designed to evaluate the impact of nucleotide changes on RNA secondary structure. However, systematic calibration of this tool for *RMRP*, which could also enable increasing its strength, could not be performed within the scope of this project.^29,30^

## Future Directions

The creation of gene-specific ACMG/AMP variant classification rules for *RMRP* is anticipated to facilitate development of specifications for other non-protein coding genes in the future. Our observations bring forth a call for increased publication of cases that bear VUS or LP *RMRP* variants *with phenotypic information as listed in Table 1*. They also demonstrate the need for clinical laboratories to obtain PP4 phenotype information from the ordering provider whenever such a variant is submitted to ClinVar. Concerningly, the VCEP experts also noted that no laboratory is routinely performing *RMRP* functional assays. The need therefore exists to promote *RMRP* functional testing to not only facilitate use of PS3 and potentially BS3, but also to upgrade their strengths with assessment of sufficient benign and pathogenic controls.^21^ Next, with continued and ongoing *RMRP* variant curation, the SCID-VCEP expects to be able to calibrate a computational prediction tool. Alternatives to RNAsnp could be tested simultaneously, including algorithms that focus upon evolutionary conservation at the *nucleotide* rather than amino acid level. Meanwhile, the SCID-VCEP will continue to explore options to modify the ACMG/AMP codes. For example, because Very Strong evidence is often necessary to classify a variant as pathogenic, but PVS1 cannot be used for *RMRP*, other pathogenic codes like PP4 may need to be thoughtfully increased in strength. It might also be possible to apply BP3 for some variants upstream of the transcription start site that do not affect the promoter, TATA box, or distance between the TATA box and transcription initiation nucleotide. Lastly, the SCID-VCEP envisions the need to convert these specifications to accommodate a fully quantitative Bayesian scoring system for variant classification.^31,32^ Such a process will necessarily require extensive testing and validation. In the meantime, these specifications will enable variant classification of numerous *RMRP* variants for clinical and research-based purposes.

## Acknowledgements

The authors thank the Clinical Genome Resource organization for feedback and support during our application. The authors also extend their gratitude to all the volunteer biocurators and expert panelists.

## The Severe Combined Immunodeficiency Disease Variant Curation Expert Panel

Additional members of the SCID-VCEP have included the following: Ariel McShane, Ben McLean, Catalin Munteanu, Joshi Stephen, Mariana Grobler, Lauren Fries, Michelle Zelnick, Neema Izadi, Pik Ying Hue, Shengfeng Xu, Wei Ma, Xinrui Shi, Xueyang Pan, Justyne Ross, Haley Garrett, Monica Sulit, Brooke Palus, and Marwa Elnagheeb.

## Data Availability

The ClinGen Severe Combined Immunodeficiency Disease Variant Curation Expert Panel submitted all variants to the ClinVar database, including evidence summaries detailing the data used for each classification. The detailed evidence used for the classification of these variants is available in the ClinGen Evidence Repository. All rules specifications are obtainable through the ClinGen Criteria Specification Registry.

## Ethics Declaration

No identifying patient information was accessed or used in this study. All data analyzed were obtained from publicly available databases, such as PubMed, and were fully anonymized before analysis. No ethical approval is required for the use of such data.

## Author Contributions

Conceptualization: I.K.C., B.A.J.; Data Curation and Formal Analysis: V.C.J., A.N-P., S.M., S.M.; Funding Acquisition: I.K.C., B.A.J.; Project Administration: I.K.C., B.A.J.; Writing-original draft: V.C.J., Y.D., A.N-P., I.K.C., B.A.J.; Writing-review and editing: V.C.J., Y.D., A.N-P., S.M., S.M., E.J.A., S.V., L.D.N., J.M.P, J.W., A.Y.C., M.J.C., M.S.H, N.S.C.v.O., J.V., B.H.Y.C., R.G., X.L., M.P., K.V.S., T.A.S., B.Y., B.A.J, I.K.C.

