## Supplementary material for "Variant Classification of a Non-Protein Coding Gene: ACMG/AMP Specifications for *RMRP* from the Clinical Genome Resource Severe Combined Immunodeficiency Disease Variant Curation Expert Panel": Table S1

| Table S1. Functional assays considered to determine pathogenicity in *RMRP* variants. | | | |
| --- | --- | --- | --- |
| **Assay** | Endonucleolytic cleavage activity | Endonucleolytic cleavage activity | Luciferase reporter assay |
| **General Description** | RNA and DNA extracted from normal human fibroblasts 12 h, 36 h, 48 h, and 60 h post-transient transfection with wild type or variant *RMRP* expression constructs were used as template for quantitative real-time PCR evaluation of *RMRP*, *CCNA2*, *CCNB2*, 5.8S rRNA, and ITS-1-bound 5.8S rRNA expression levels | RNA and DNA extracted from normal human fibroblasts transiently transfected with wild type or variant *RMRP* expression constructs were used as template for quantitative real-time PCR evaluation of *RMRP*, *CCNB2*, 5.8S rRNA, and ITS-1-bound 5.8S rRNA expression levels | Expression plasmids containing wild type and variant *RMRP* promoter sequences upstream of an shRNA against luciferase were transfected in COS7 cells in combination with a luciferase expression plasmid. |
| **Cells** | Normal human fibroblasts transiently transfected with wild type or variant *RMRP* expression constructs | Normal human fibroblasts transiently transfected with wild type or variant *RMRP* expression constructs | COS7 cells transfected with a luciferase-targeted shRNA under the control of wild type or variant *RMRP* promoters |
| **Output** | Cyclin levels (based on *CCNA2* and *CCNB2* mRNA levels); rRNA cleavage activity normalized to wild type (ratio of levels of cleaved to uncleaved 5.8S rRNA) | mRNA cleavage activity normalized to wild type (inverse relative increase of *CCNB2* mRNA level); rRNA cleavage activity normalized to wild type (ratio of levels of cleaved to uncleaved 5.8S rRNA) | Relative luciferase activity (promoter strength correlates with the degree of shRNA-mediated downregulation of luciferase expression) |
| **Variants tested** | Cartilage-hair hypoplasia:  NR_003051.3:n.71A>G (NC_000009.12:g.35657948T>C)  Anauxetic dysplasia:  NR_003051.3:n.15G>A (NC_000009.12:g.35658004C>T)  NR_003051.3:n.112_113insACGTAGACATTCCT (NC_000009.12:g.35657906_35657907insAGGAATGTCTACGT)  NR_003051.3:n.91_92AG>GC (NC_000009.12:g.35657927_35657928delinsGC)  NR_003051.3:n.255G>C (NC_000009.12:g.35657764G>C) | Cartilage-hair hypoplasia:  NR_003051.3:n.64C>T (NC_000009.12:g.35657955G>A)  NR_003051.3:n.97_98dup (NC_000009.12:g.35657922_35657923dup)  NR_003051.3:n.71A>G (NC_000009.12:g.35657948T>C)  NR_003051.3:n.127C>T (NC_000009.12:g.35657892G>A)  NR_003051.3:n.147G>A (NC_000009.12:g.35657872C>T)  NR_003051.3:n.5C>T (NC_000009.12:g.35658014G>A)  NR_003051.3:n.221T>C (NC_000009.12:g.35657798A>G)  NR_003051.3:n.249C>T (NC_000009.12:g.35657770G>A)  NR_003051.3:n.262C>T NC_000009.12:g.35657757G>A  NR_003051.3:n.196C>T (NC_000009.12:g.35657823G>A)  Anauxetic dysplasia:  NR_003051.3:n.112_113insACGTAGACATTCCT (NC_000009.12:g.35657906_35657907insAGGAATGTCTACGT)  NR_003051.3:n.91_92AG>GC (NC_000009.12:g.35657927_35657928delinsGC)  NR_003051.3:n.255C>G (NC_000009.12:g.35657764G>C) | All cartilage-hair hypoplasia:  NR_003051.3:n.90C>G (NC_000009.12:g.35657929G>C)  NR_003051.3:n.125C>T (NC_000009.12:g.35657894G>A)  NR_003051.3:n.71A>G (NC_000009.12:g.35657948T>C)  NR_003051.3:n.-13_-24dup (NC_000009.12:g.35658032_35658041dup)  NR_003051.3:n.181G>A (NC_000009.12:g.35657838C>T) |
| **Reference**  **(PMID)** | Thiel et al., 2005. (16252239) | Thiel et al., 2007. (17701897) | Hermanns et al., 2005 (16254002) |
