## Supplementary material for "Variant Classification of a Non-Protein Coding Gene: ACMG/AMP Specifications for *RMRP* from the Clinical Genome Resource Severe Combined Immunodeficiency Disease Variant Curation Expert Panel": Table S2

Table S2. Point-based assessment of *de novo* occurrences according to phenotypic specificity

| **Phenotypic consistency** | **Points per proband** | |
| --- | --- | --- |
|  | **Confirmed *de novo***  **(PS2)^a^** | **Assumed *de novo***  **(PM6)^b^** |
| Phenotype highly specific for gene (PP4_Moderate) | 2 | 1 |
| Phenotype consistent with gene but not highly specific (PP4) | 1 | 0.5 |
| Phenotype consistent with gene but not highly specific and with high heterogeneity ^c^  (PP4 not met but greater than zero phenotype points) | 0.5 | 0.25 |
| Phenotype not consistent with gene | No points awarded | |

^a^ *de novo* with confirmed parental relationships. ^b^ *de novo* with unconfirmed parental relationships  ^c^ Maximum allowable value of 1 may contribute to the overall score
